# The role of systemic therapies in current and emerging opportunities for de-intensification in melanoma: a scoping review protocol

**DOI:** 10.1101/2023.11.21.23298799

**Authors:** Jennifer A Soon, Fanny Franchini, Maarten Ijzerman, Grant A McArthur

**Affiliations:** Sir Peter MacCallum Department of Oncology, University of Melbourne, Australia; Cancer Health Services Research, University of Melbourne, Australia; Peter MacCallum Cancer Centre, Melbourne, Australia; Erasmus School of Health Policy & Management, Rotterdam, The Netherlands; Victorian Comprehensive Cancer Centre Alliance, Melbourne, Australia

**Keywords:** de-intensification, de-escalation, melanoma, over-treatment, low value care

## Abstract

**Objective:** The purpose of this scoping review is to identify the role of systemic therapies in current and emerging opportunities to de-intensify systemic treatment of cutaneous melanoma. It will also seek to comment on the proportion of studies that include patient-reported outcomes and quality of life measures.

**Introduction:** With healthcare costs rising, focus is shifting towards maximising health outcomes per dollar. One approach to optimising value is through de-intensification, which is the rationalisation of routine treatment without compromising patient outcomes. Since 2013, successful clinical trials of high-cost immune checkpoint, BRAF and MEK inhibition have led to these drugs becoming ubiquitous in melanoma management. Pipeline therapies such as relatlimab and Tumour Infiltrating Lymphocyte (TIL) therapy are expected to have a similar or even greater cost. We hypothesize that neoadjuvant and response-directed strategies will be identified as well as the emerging potential of prognostic and predictive tools.

**Inclusion criteria:** This review aims to include studies of adult patients with cutaneous melanoma that report on de-escalation or de-intensification of internationally accepted standard-of-care systemic therapies or the use of systemic therapies to de-intensify subsequent surgery or radiotherapy. Systemic treatment across any stage of disease will be considered. A full-text English-language version must be available for a study to be eligible.

**Methods:** A systematic search strategy has been developed for MEDLINE, EMBASE and PubMed from 1 January 2013 to 30 June 2023. Additional texts will be sourced from grey literature, Google Scholar and reference scanning. Two authors will screen abstracts and full texts facilitated by the Covidence software. Disagreements will be resolved by consensus or a third reviewer. The first author (JS) will perform data extraction whilst a second author (FF) will review a random selection of papers to ensure consistent interpretation. De-intensification strategies will be categorised by concept, potential impact on resource utilisation and patient outcomes, and strength of evidence. Data will be synthesised qualitatively and quantitatively. Results will be reported following the Preferred Reporting Items for Systematic Reviews and Meta-Analyses extension for Scoping Reviews (PRISMA-ScR).

The results of this scoping review will directly inform a melanoma consumer and clinician survey exploring their perspectives on de-intensification strategies.

## Introduction

The last decade has witnessed a truly impressive reversal in prospects for patients with advanced melanoma. Previously, systemic options for advanced melanoma were limited to chemotherapy, which resulted in a median overall survival benefit of six to ten months (1). However, in 2011 the United States Food and Drug Administration (US FDA) approved two novel medicines that heralded a paradigm shift in the treatment of advanced melanoma: ipilimumab, a Cytotoxic T-Lymphocyte Antigen-4 (CTLA-4) checkpoint inhibitor (CPI) of the immune system, and vemurafenib, a small molecule kinase inhibitor effective in *BRAF*-mutant melanoma. In the following years, these drugs were superseded by more effective first-line agents and combinations. Pembrolizumab and nivolumab, anti-Programmed-Death-1 CPI, were both FDA-approved in 2014, and demonstrated greater efficacy with far less toxicity. The FDA approved the current gold standard treatment, ipilimumab in combination with nivolumab, in 2015, which demonstrates an impressive 48.5% survival rate at 7.5 years of follow-up (2). For the first time in history, patients with advanced melanoma had a chance of long-term survival.

Immunotherapies and targeted therapies were quickly moved into the adjuvant setting to reduce the risk of relapse for resected stage III melanoma, that is melanoma with nodal involvement or in-transit disease that has been completely excised with surgery. An overall survival benefit has been demonstrated with ipilimumab compared to placebo, although the uptake of this treatment has been limited by the 41.6% grade 3/4 immune-related adverse event rate (3). Pembrolizumab, nivolumab and the combination of dabrafenib and trametinib have shown a statistically significant relapse-free survival benefit, however data is not yet mature enough to demonstrate an overall survival benefit (4–6). More recently, the FDA has approved pembrolizumab for resected high-risk stage II melanoma, and promising clinical trials are underway to establish the role of neoadjuvant CPI for stage III melanoma (7). However, these medicines do come at a significant cost to healthcare payers and can be associated with permanent and life-changing toxicities. Since overall survival benefit is not yet established in adjuvant therapy, questions are being raised in melanoma around how much treatment is required and whether we are over-treating patients.

De-intensification is the reduction of intensity or duration of treatment without compromising patients’ cancer outcomes (8). Increased understanding around the biology of molecular and genomic subtypes of cancer are allowing clinicians to identify patients who stand to gain only marginal benefits from standard-of-care (SOC) treatment. For patients, de-escalating treatment offers an opportunity to avoid physical, financial and time toxicities with resulting improvement in quality of life (QoL). For society, the implications of de-intensification include a reduction in health resource utilisation, allowing surplus resources to be diverted to other endeavours offering greater benefit. This concept of de-intensification is more mature in the disciplines of breast, head and neck, and genitourinary oncology. De-intensification approaches have been explored in breast oncology since the early 2000s (9) and significant progress has been made to de-intensify in oropharyngeal squamous cell carcinomas associated with human papilloma virus (10). However, integration into SOC guidelines has been hampered by the lack of quality randomised data to support de-intensification approaches (11). As therapeutic options for cancer patients improve and survival gains become incremental, there is growing recognition of the need for more clinical trials addressing this issue. Beyond this, there is the need to standardise or establish minimum data requirements to ensure the quality of data is synthesizable and sufficient to support practice change (8,12).

In melanoma, de-intensification of treatment has been integrated mainly into the surgical pathways. Externally validated predictive nomograms (13,14) estimating the risk of sentinel lymph node positivity can differentiate high from low-risk patients with stage I or II melanoma. In low-risk patients, omission of sentinel lymph node biopsy can be considered, which saves patients from a low-yield biopsy. The MSLT-II study (15) has driven surgical de-intensification by providing randomised data showing no difference in melanoma-specific survival in sentinel-node positive patients who undergo observation rather than immediate complete lymph node dissection. As such, SOC has now shifted away from these procedures, which are often associated with surgical morbidity and poorer QoL. However, de-intensification of systemic therapies remains a relatively novel concept in melanoma that has not yet been translated into clinical practice. Given the toxicity profile of systemic therapies for melanoma and their high cost, exploring safe de-intensification represents a promising opportunity to improve patients’ experiences and outcomes whilst also reducing healthcare resource utilisation.

Currently, de-intensification approaches are not systematically described in melanoma: a preliminary search of MEDLINE, PROSPERO, the Cochrane Database of Systematic Reviews and JBI Evidence Synthesis was unable to identify any published or in progress systematic or scoping reviews on this topic. Therefore, the purpose of this scoping review is to systematically describe the current and emerging opportunities to de-intensify systemic treatment in melanoma. This overview will more definitively outline the knowledge gaps and identify potential high-value prospects in de-intensification that can guide future research.

## Review questions

1. What is the current role of systemic therapies in de-intensifying treatment of melanoma?
2. What is the role of systemic therapies in emerging opportunities to de-intensify treatment of melanoma?
3. What proportion of these studies include patient-reported outcomes or quality of life measures?
4. What is the anticipated impact of de-intensification strategies on clinical outcomes, patient-reported outcomes, quality of life measures and other metrics such as financial and time toxicity?

## Eligibility criteria

### Population

Eligible studies will involve adult patients with cutaneous melanoma at any stage of disease. Studies looking exclusively at non-cutaneous melanoma, for example mucosal or uveal, will not be included.

### Concept

This review will focus on the role of systemic therapies in de-intensification of internationally accepted SOC, specifically identifying opportunities to either de-intensify systemic therapy or use systemic therapy to de-intensify radiotherapy or surgery. SOC is defined by adjuvant CPI (anti-PD-1) and targeted therapies in resected stage III/IV disease, and CPI (anti-CTLA-4, anti-PD-1, anti-LAG-3) and targeted therapies in advanced melanoma.

Studies of risk prediction tools or biomarkers that can aid in treatment de-intensification will also be included. Studies should contain at least 25 melanoma patients or samples. In the context of treatment de-intensification, biomarkers for surveillance, disease monitoring and prediction of therapeutic response are the most pertinent. A predictive biomarker is an indicator of the likely treatment benefit for a patient, whereas a prognostic biomarker “provides information on the likely health outcome… irrespective of treatment” (16). For a biomarker to be considered predictive, its presence must be associated with a difference in treatment effect. A prognostic biomarker may be used in a predictive model (e.g., a nomogram) but may not necessarily be a predictive biomarker on its own. A purely prognostic biomarker has an additional evidentiary step to demonstrate its predictive potential and that it can safely guide treatment decisions – for this reason, prognostic biomarkers will not be included in this scoping review. Predictive biomarkers will only be considered if they are cross validated and demonstrate an association with an endpoint of interest, for example treatment response, or likelihood of relapse. Studies of unvalidated biomarkers and prediction tools will be excluded, as will diagnostic biomarkers and those predicting treatment toxicity only.

### Context

This review will consider studies where internationally accepted SOC treatment for melanoma is readily accessible by the general population. Studies conducted exclusively in a resource-poor setting will not be included due to the differences in SOC management. There are no limitations on healthcare settings or geographic locations.

### Types of sources

This scoping review will consider a multitude of trial designs including, but not limited to, randomized controlled trials, non-randomized controlled trials, before and after studies and interrupted time-series studies. In addition, analytical observational studies including prospective and retrospective cohort studies, case-control studies and analytical cross-sectional studies will be considered for inclusion. This review will also consider descriptive observational study designs and descriptive cross-sectional studies for inclusion. Individual case reports and case series lie outside the scope of this review as their management may not be applicable to wider populations nor reflective of broader trends. Opinion papers, narrative reviews and other grey literature will also be considered for inclusion in this scoping review.

## Methods

The scoping review will be conducted in accordance with the JBI methodology for scoping reviews.

### Search strategy

A preliminary search of MEDLINE and Google Scholar was undertaken to identify articles on the topic. The text words contained in the titles and abstracts of relevant articles, and the index terms used to describe the articles were used to develop a full search strategy for MEDLINE, EMBASE, and Google Scholar (Appendix 1a, 1b). The search strategy, including all identified keywords and index terms, will be adapted for each included database and information source. Reference lists from included studies will be scanned for any additional relevant texts.

Only full-text studies published in English will be included. Studies published between 1 January 2013 to 30 June 2023 will be included as the first modern immunotherapies and targeted therapies in melanoma were only approved by the United States’ Food and Drug Administration (FDA) in late 2011, and hence uptake of these medicines was not fully established until 2013 and beyond. De-intensification strategies are unlikely to have been trialled or reported on prior to this date.

### Study selection

Texts and sources will be uploaded into the Covidence software for reference management and duplicates removed. Initial pilot testing will be conducted to ensure screening inclusion criteria are appropriate. Subsequently, titles and abstracts will be screened by two independent reviewers against the screening inclusion criteria. Full-text screening by two or three independent reviewers will allow further refinement of the included sources based on a more detailed assessment against the inclusion criteria. Exclusion reasons for all sources will be reported. Disagreements will be resolved by consensus, or failing that, the involvement of a third reviewer.

The results of the search and the study inclusion process will be reported in full in the final scoping review and presented in a Preferred Reporting Items for Systematic Reviews and Meta-analyses extension for Scoping Review (PRISMA-ScR) flow diagram (17).

### Data extraction

A data extraction tool will be designed and piloted based on the key outputs required to answer the research questions. The extraction tool will be tailored to the study type as demonstrated in Figure 1. For instance, commentaries and non-systematic reviews will undergo a truncated data extraction tool (“Section B”), and studies reporting biomarkers in the early phases of development will go through short-form data extraction tool with specific biomarker-related fields (“Section C”). All other studies will be analysed using a comprehensive data extraction tool (“Section A”). A draft extraction tool showing all sections can be reviewed in Appendix 2. We will take an iterative approach to the development of the data extraction tool: modifications may be required as patterns and trends emerge, and the rationale behind the changes will be detailed in the scoping review. The data extracted will include specific details about the participants, concept, context, study methods, potential impact on resource utilisation and patient outcomes, and strength of evidence. A companion extraction information sheet will be developed to minimise variation in the application of the data extraction tool.

**Figure 1:**
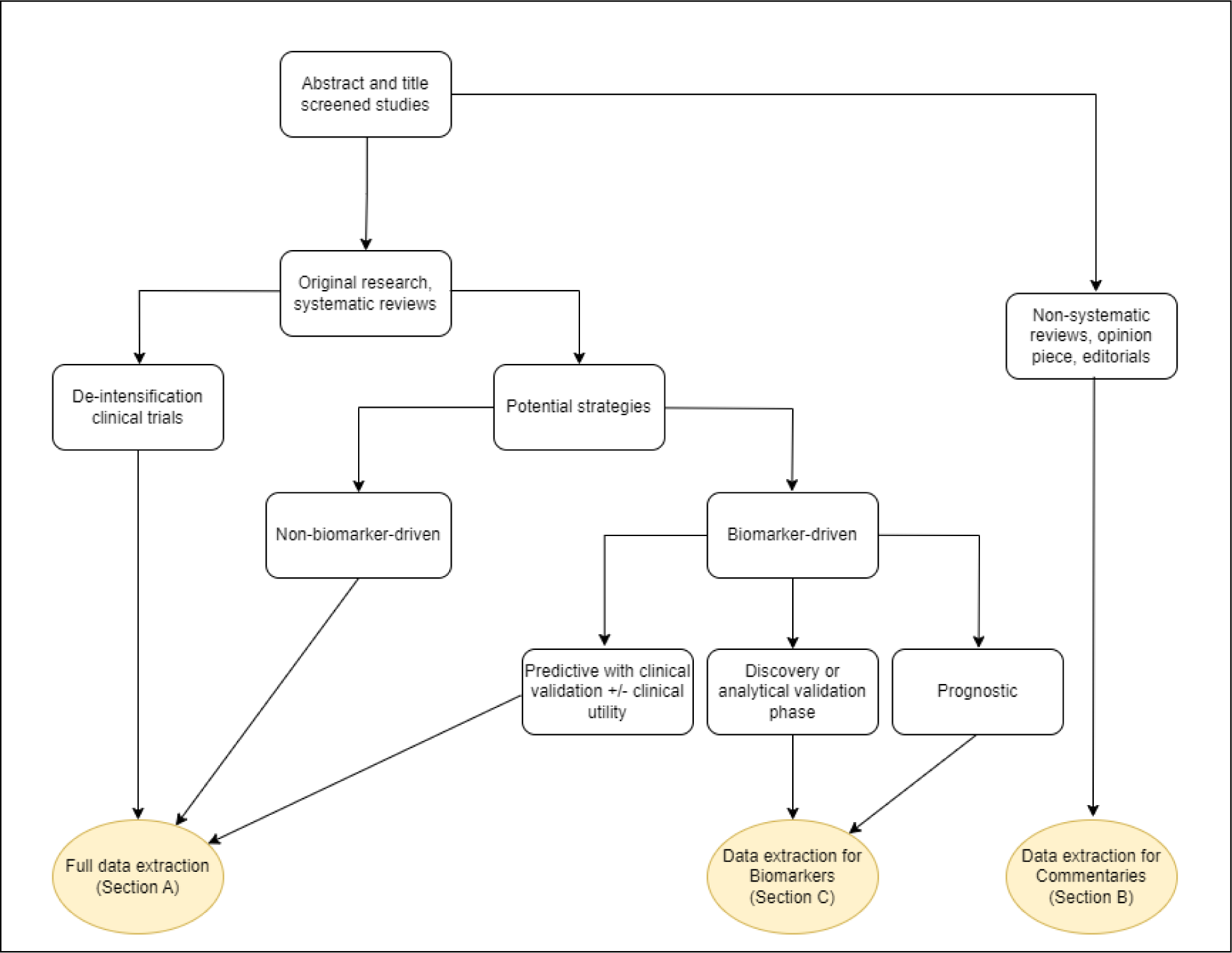
Conceptual representation of which studies will undergo specific sections of the extraction tool.

Data extraction will be performed by a primary reviewer. A secondary reviewer will assess a random selection of ten percent (or at least twenty) of the eligible full texts to ensure consistency in application of the data extraction tool. Any disagreements that arise between the reviewers will be resolved through discussion or, where consensus is not possible, after discussion with a third reviewer. Formal assessment of risk of bias lies outside the remit of a scoping review.

### Data analysis and presentation

Data will be grouped into categories of de-intensification based on approach and purpose. Descriptive statistics will be used to summarise the extracted data, which will also be presented graphically using visualisation techniques such as concept mapping.

## Ethics

This scoping review protocol, focusing on de-intensification in melanoma, exclusively involves the synthesis of published works and does not involve the collection or analysis of data from human participants. Therefore, formal ethics approval was not required. The study adheres to established ethical guidelines, ensuring confidentiality and responsible research conduct. All efforts have been made to uphold transparency and ethical standards throughout the scoping review process. For any inquiries concerning ethical considerations, please contact the corresponding author, Dr Jennifer Soon.

## Data Availability

All data produced in the present work are contained in the manuscript.

## Acknowledgements

We wish to acknowledge the generous expertise of Ms Smaro Lazarakis, Clinical and Research Librarian at the Health Sciences Library of Melbourne Health, in developing the database search strategy.

This scoping review will be conducted as part of Dr Jennifer Soon’s doctorate studies. She acknowledges the stipend support she receives from the University of Melbourne through the Melbourne Research Scholarship.

## Appendices

### Appendix 1a: Proposed search strategy – MEDLINE, EMBASE

| Concept | Search terms |
| --- | --- |
| **Melanoma** | exp melanoma/dt<br>OR<br>exp melanoma/ and exp antineoplastic agents<br>OR<br>melanoma*.tw,kf. |
| **Systemic therapies** | (immunotherap* or pembrolizumab or nivolumab or ipilimumab or dabrafenib or trametinib or vemurafenib or cobimetinib or encorafenib or binimetinib or anti-PD* or PD-1 antibod* or anti-CTLA or CTLA-4 or BRAF inhibitor* or MEK inhibitor* or lag-3 or relatlimab or lag inhibitor* or drug treatment* or drug therap* or systemic treatment* or systemic therap* or pharmacotherap* or antineoplastic* or targeted therap* or adjuvant treatment* or adjuvant therap*).tw,kf. |
| **De-intensification** | exp medical overuse/ or exp inappropriate prescribing/ or exp deprescriptions/ or exp low-value care/<br>OR<br>(low value or de-implement* or unnecessary prescri* or overuse or overutili* or overtreat* or de-escalat* or de-prescri* or de-intensif* or discontinu* or undertreat* or cessation or response-directed or de-implement* or deimplement* or over prescri* or deprescri* or inappropriate prescri*).tw,kf.<br>OR<br>((predict* or prognos*) adj5 (model* or tool* or instrument* or algorithm or test* or measur*)).tw,kf. |
| **Limits** | Human studies<br>English language<br>1<sup>st</sup> January 2013 to 30<sup>th</sup> June 2023 |

### Appendix 1b: Proposed search strategy – Google Scholar

Search terms: melanoma de-intensification OR discontinuation OR discontinue OR de-intensify OR “guide treatment” OR “predict treatment response” OR “response-directed”

First 200 studies will be reviewed

### Appendix 2: Data extraction tool

Updated 18th October 2023

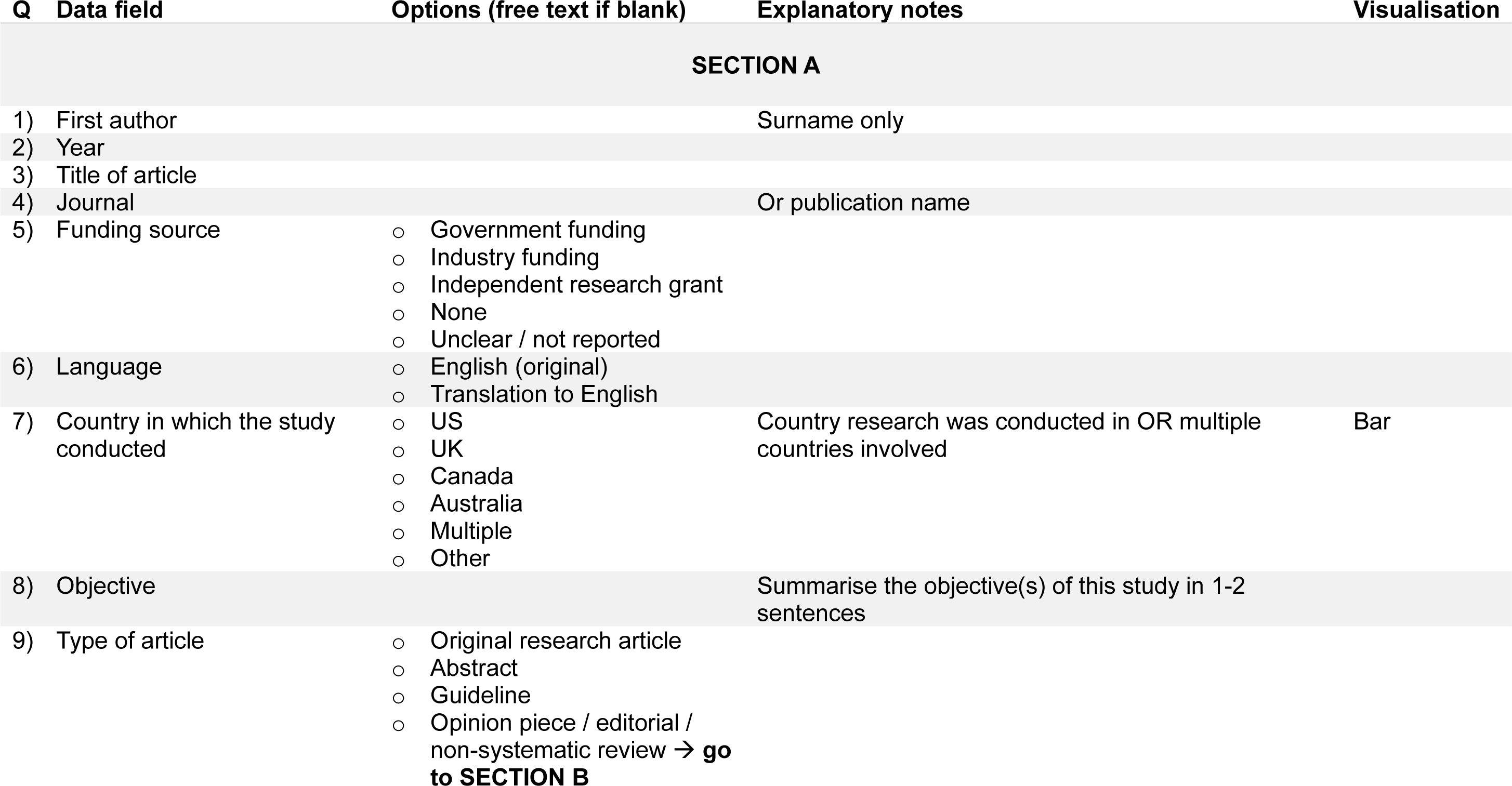
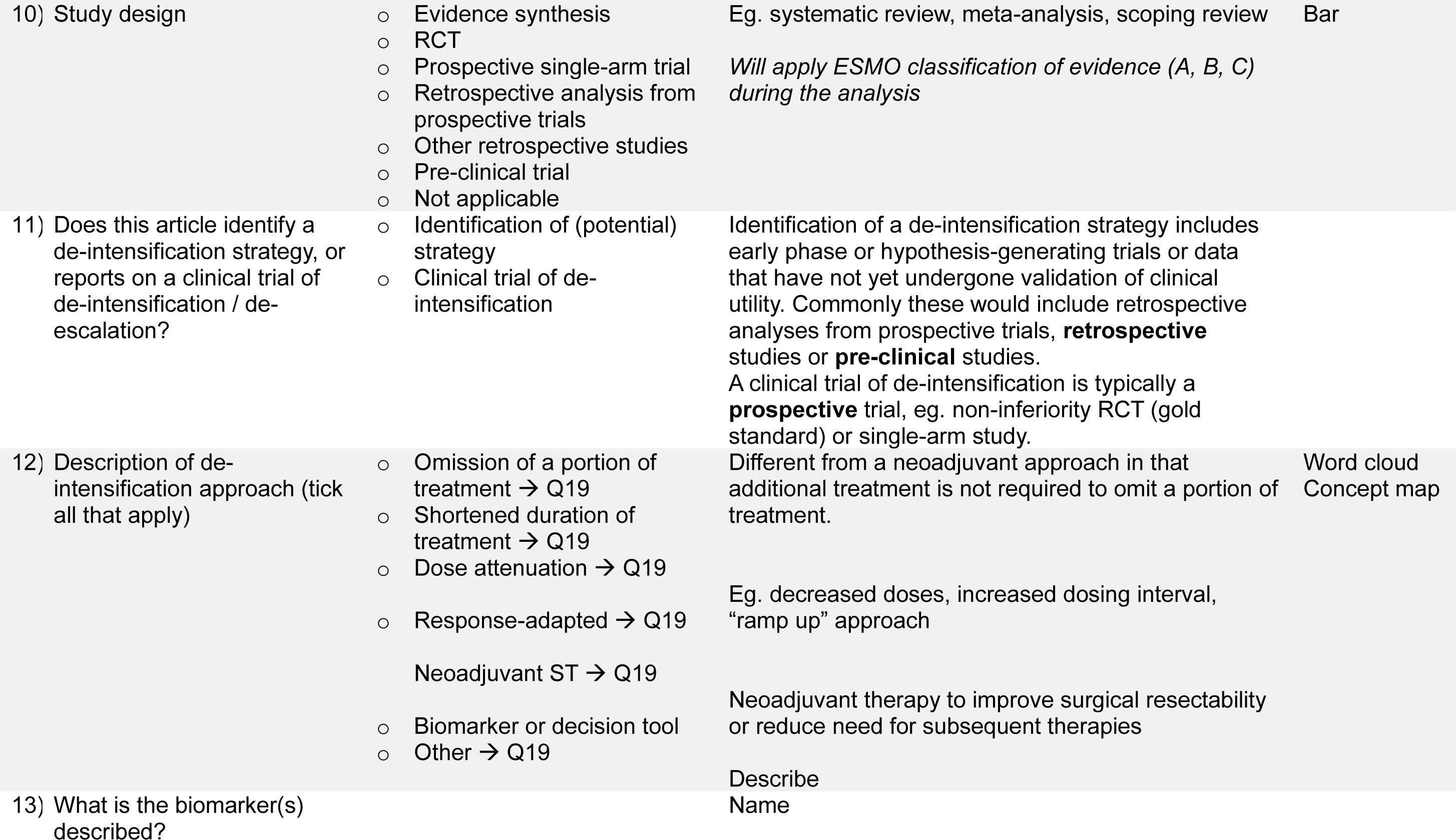
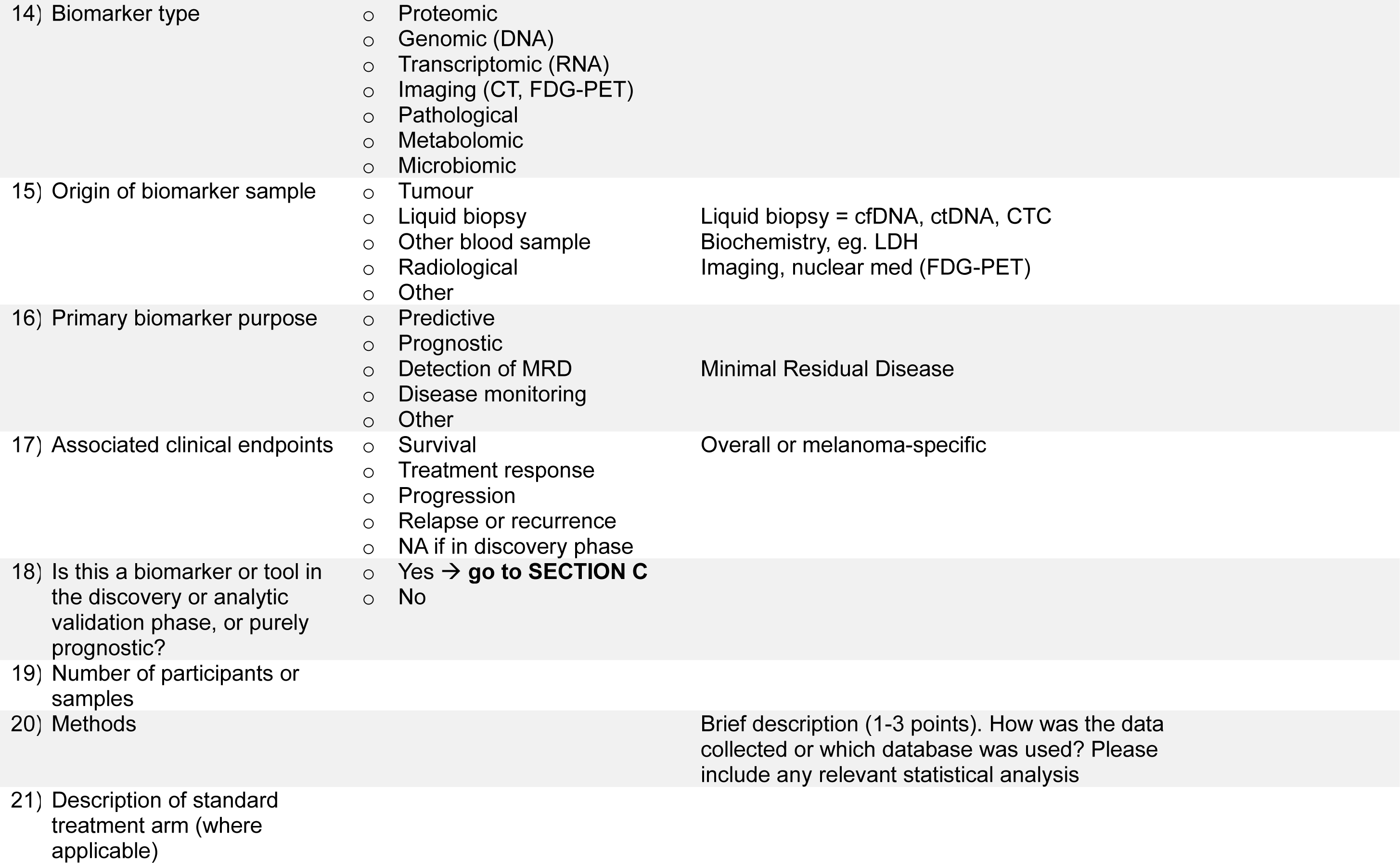
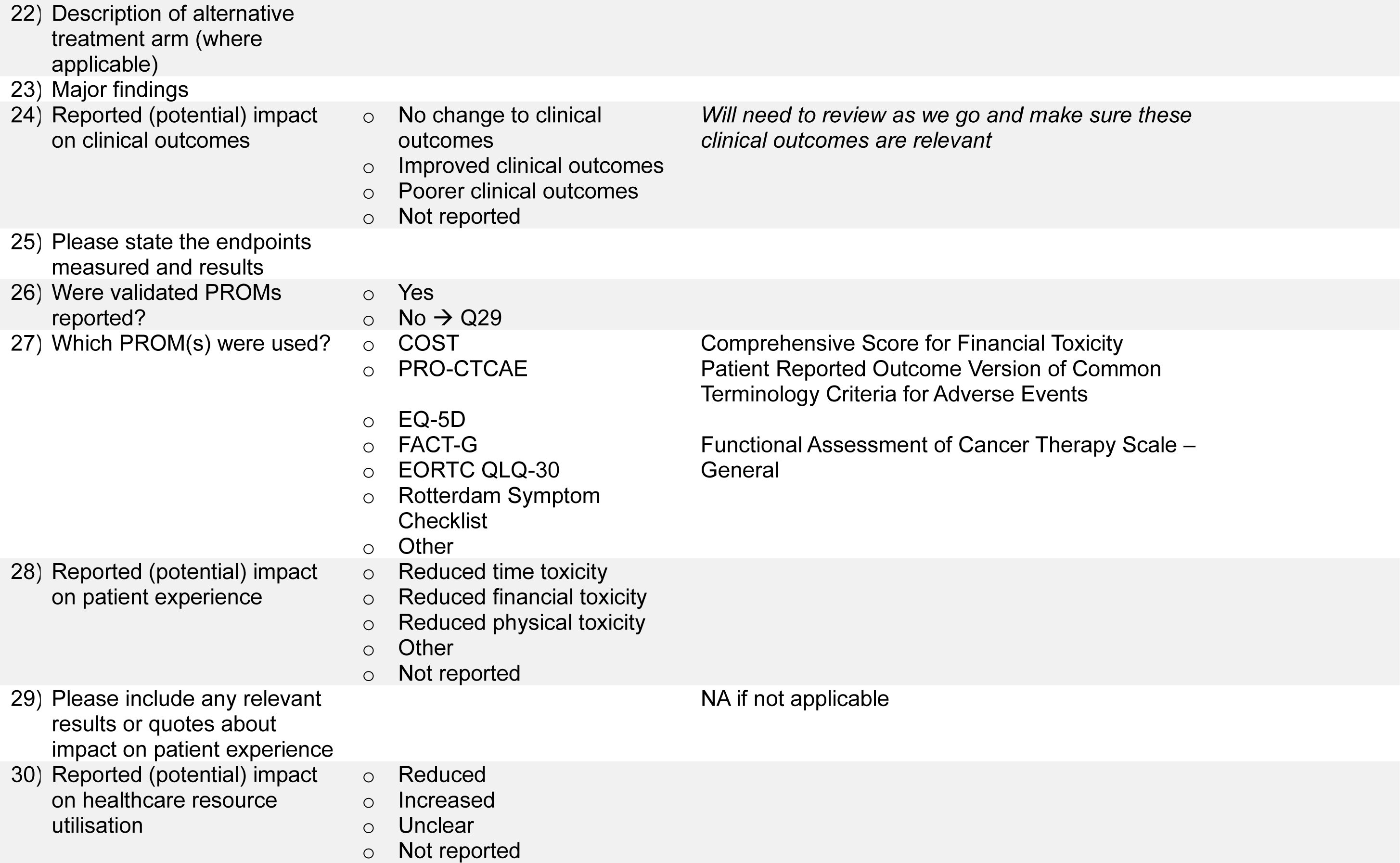
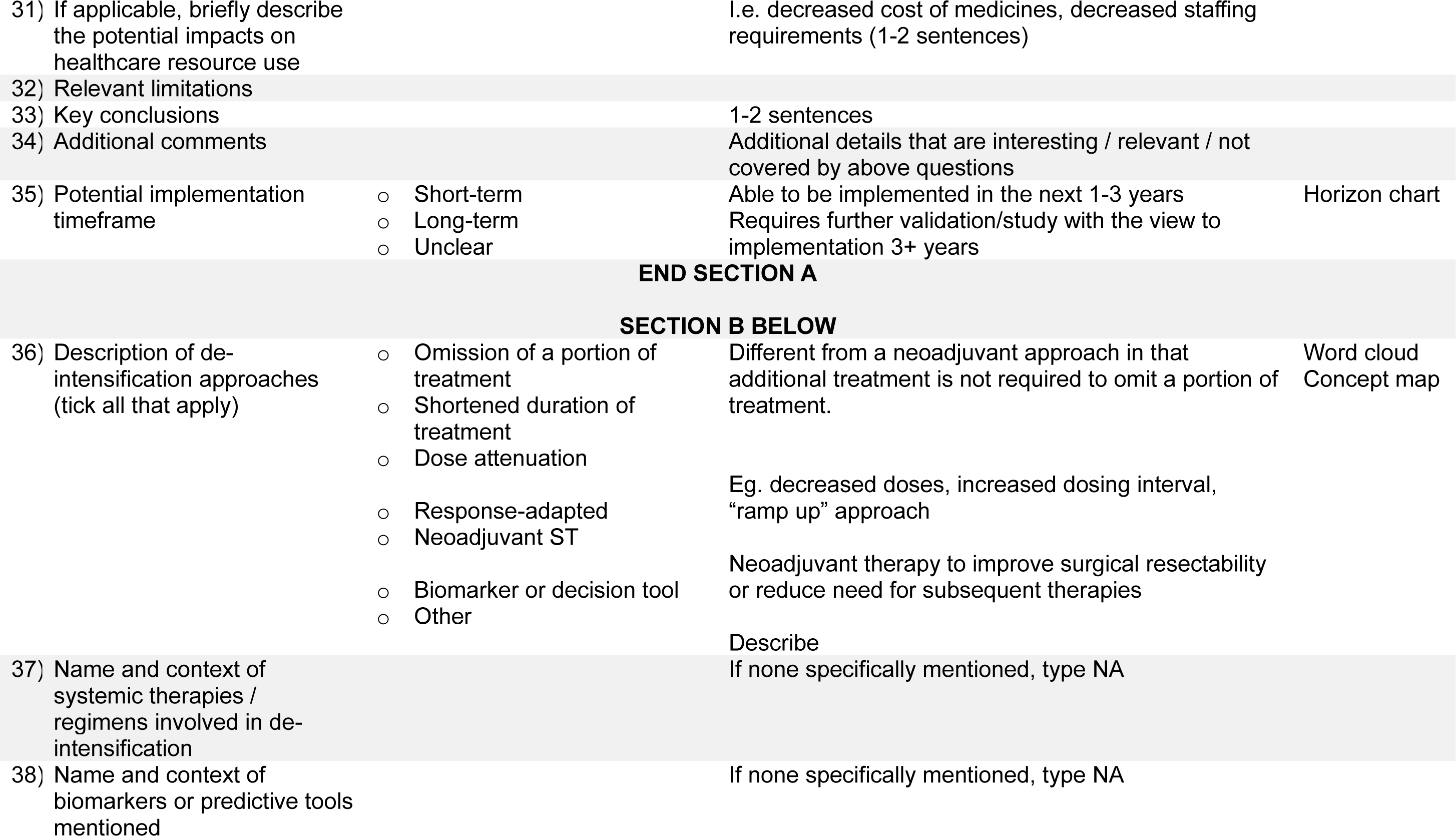
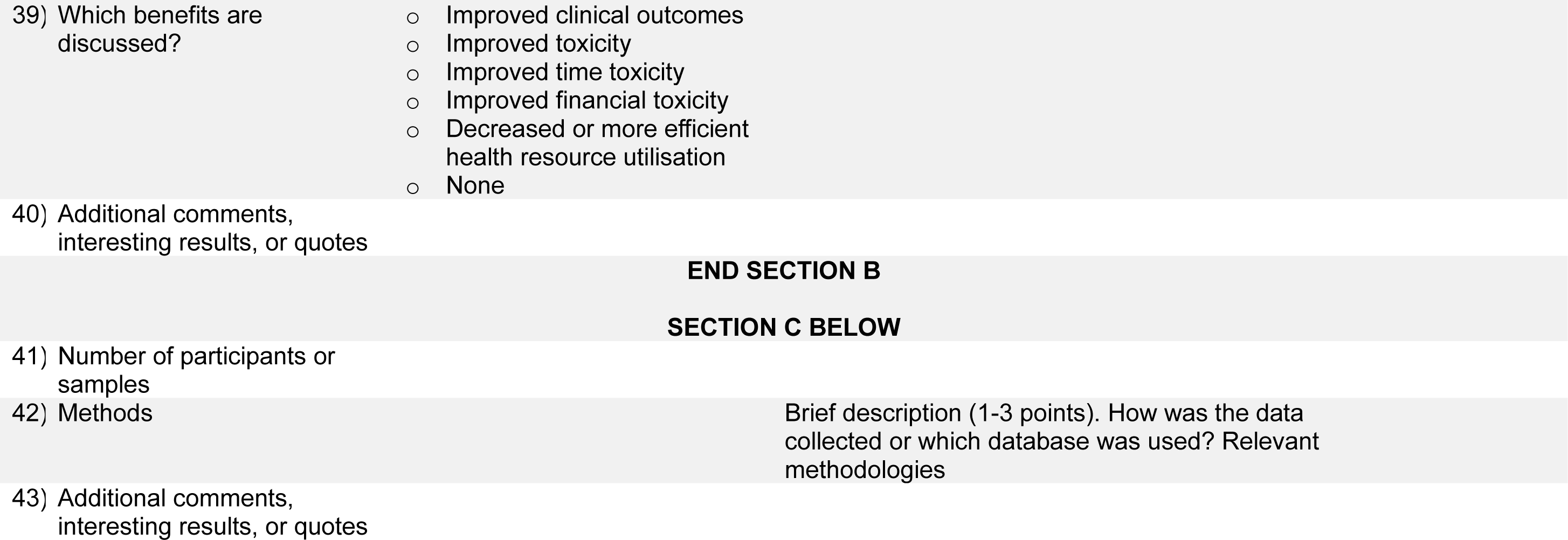

